# Alcohol use and cognitive functioning in young adults: improving causal inference

**DOI:** 10.1101/19003327

**Authors:** Liam Mahedy, Steph Suddell, Caroline Skirrow, Gwen S. Fernandes, Matt Field, Jon Heron, Matthew Hickman, Robyn Wootton, Marcus R. Munafò

## Abstract

**Background and Aims:** There have been few longitudinal studies of association between alcohol use and cognitive functioning in young people. We aimed to examine whether alcohol use is a causal risk factor for deficient cognitive functioning in young adults.

**Design:** Linear regression was used to examine the relationship between longitudinal latent class patterns of binge drinking and subsequent cognitive functioning. Two-sample Mendelian randomisation (MR) tested evidence for the causal relationship between alcohol use and cognitive functioning.

**Setting:** South West England.

**Participants:** The observational study included 3,155 adolescents and their parents (fully adjusted models) from the Avon Longitudinal Study of Parents and Children (ALSPAC). Genetic instruments for alcohol use were based on almost 1,000,000 individuals from the GWAS & Sequencing Consortium of Alcohol and Nicotine use (GSCAN). Genome-wide association studies for cognitive outcomes were based on 2,500 individuals from ALSPAC.

**Measurements:** Binge drinking was assessed at approximately 16, 17, 18, 21, and 23 years. Cognitive functioning comprised working memory, response inhibition, and emotion recognition assessed at 24 years of age. Ninety-nine independent genome-wide significant SNPs associated with ‘number of drinks per week’ were used as the genetic instrument for alcohol consumption. Potential confounders were included in the observational analyses.

**Findings:** Four binge drinking classes were identified: ‘low-risk’ (41%), ‘early-onset monthly’ (19%), ‘adult frequent’ (23%), and ‘early-onset frequent’ (17%). The association between early-onset frequent binge drinking and cognitive functioning: working memory (*b*=0.09, 95%CI=-0.10 to 0.28), response inhibition (*b*=0.70, 95%CI=-10.55 to 11.95), and emotion recognition (*b*=0.01, 95%CI=-0.01 to 0.02) in comparison to low-risk drinkers were inconclusive as to whether a difference was present. Two-sample MR analyses similarly provided little evidence that alcohol use is associated with deficits in working memory using the inverse variance weight (*b*=0.29, 95%CI=-0.42 to 0.99), response inhibition (*b*=-0.32, 95%CI=-1.04 to 0.39), and emotion recognition (*b*=0.03, 95%CI=-0.55 to 0.61).

**Conclusions:** Binge drinking in adolescence and early adulthood may not be causally related to deficiencies in working memory, response inhibition, or emotion recognition in youths.

## INTRODUCTION

Alcohol use during adolescence, when the brain is still developing and undergoing considerable structural and functional changes (1) is a major public health concern. The association between binge drinking and cognitive functioning (i.e., working memory, response inhibition, and emotion recognition) has received particular attention because some cognitive functions do not peak until early adulthood (2–5) in parallel with maturation of the prefrontal cortex (6,7).

There is a wealth of evidence from animal (8,9), neuroimaging (10–12), twin (13,14), and cognitive neuroscience (15,16) studies suggesting that adolescent binge drinking is negatively associated with cognitive functioning. However, the direction of this association remains unclear as many of these results are based on evidence from small cross-sectional studies. Studies that have examined this association using prospective data have largely revealed mixed findings. For example, some studies have found that alcohol use preceded deficits in domains of cognitive functioning (17–20), while other studies have provided support for the opposite direction (21–23). One possible way to overcome reverse causation is to use Mendelian randomisation (24). This approach uses genetic variants to proxy for an exposure in an instrumental variable analysis to estimate the causal effect on the outcome (25). One previous study examining the association between alcohol use and cognition in mid-to late-adulthood using observational and MR approaches (26), found that having consumed ‘any versus no’ alcohol was associated with better immediate recall, delayed recall, verbal fluency, and processing speed in the observational study, however these findings were not supported by the MR analyses.

In an effort to strengthen the evidence we used a triangulation approach with observational and genetic epidemiological methods to better understand the causal relationships between drinking patterns and cognitive functioning in young adults using data from Avon Longitudinal Study of Parents and Children (ALSPAC). The aims were to investigate

1. whether patterns of binge drinking (assessed between 16 to 23 years) were associated with working memory, response inhibition, and emotion recognition assessed at age 24, and
2. whether alcohol use was associated with cognitive functioning using two-sample Mendelian randomisation (MR) (27). MR can reduce bias from residual confounding and reverse causation by using genetic variants that are known to be associated with the exposure (25). We expected to find that more frequent binge drinking would be associated with deficient cognitive functioning, and that this association would be supported by the MR analyses.

## METHODS

### Design

Longitudinal latent class analysis was used to derive heterogenous patterns of binge drinking from ages 16 to 23 years. Linear regression was used to examine the relationship between patterns of binge drinking and subsequent cognitive functioning. The young person provided self-reported information on binge drinking and cognitive functioning. The clear temporal ordering of exposure, confounders and outcome helps to rule out the possibility of reverse causality. Two-sample MR tested evidence for the causal relationship between alcohol use and cognitive functioning.

## Observational analyses

### Participants and Procedure

We used data from the Avon Longitudinal Study of Parents and Children (ALSPAC), an ongoing population-based study that contains a wide range of phenotypic and environmental measures, genetic information and linkage to health and administrative records. A fully searchable data dictionary is available on the study’s website http://www.bristol.ac.uk/alspac/researchers/our-data/. Approval for the study was obtained from the ALSPAC Ethics and Law Committee and the Local Research Ethics Committees. Informed consent for the use of data collected via questionnaires and clinics was obtained from participants following recommendations of the ALSPAC Ethics and Law Committee at the time. Consent for biological samples has been collected in accordance with the Human Tissue Act (2004). All pregnant women residing in the former Avon Health Authority in the south-west of England and had an estimated date of delivery between 1 April 1991 and December 1992 were eligible for the study (Phase I consisted of *N*=14,541). Of the 13,988 offspring alive at one year, a small number of participants have withdrawn fully from the study (*n*=41), leaving an eligible sample of 13,947. Of these, 9,299 offspring were invited to attend the 24-year clinic assessment. Detailed information about ALSPAC is available online www.bris.ac.uk/alspac and in the cohort profiles (28–30). A detailed overview of our study population, including attrition at the different measurement occasions is presented in Supplementary Material Figure S1.

## Measures

A timeline of data collection is presented in Supplementary Material Figure S2.

### Exposure: Binge drinking

Information on binge drinking was collected on five occasions via a questionnaire (Q) or during attendance at a study clinic (C). Mean ages at response were: 16y 7m (Q), 17y 9m (C), 18y 6m (Q) 20y 11m (Q), and 22y 11m (Q) using the following question reflecting drinking over the past year “How often do you have six or more drinks on one occasion?”. One drink was specified as ½ pint (568 ml) average strength beer/lager, one glass of wine, or one single measure (25ml) of spirits. Responses at each timepoint were used to derive a repeated 3 level ordinal variable with categories “Never/Occasional” (comprising of “Never” and “Less than monthly”), “Monthly” and “Weekly”. Daily or almost daily was collapsed into the “Weekly” group.

### Outcome variables

At 24 years of age (*M*=24.0 years; *SD*=9.8 months) participants attended a clinic-based assessment which included computerised cognitive assessments as part of a broader assessment battery of mental and physical health and behaviour. Data collection for the online questionnaires was collected and manged by REDcap electronic data capture tools (31,32).

### Working memory

The *N*-back task (2-back condition) was used to assess working memory. The *N*-back task (33) is widely used to measure working memory (17,34,35). A measure of discriminability (*d*′) was chosen as the primary outcome measure given it is an overall performance estimate. Of the participants assessed with cognitive tasks at age 24 (*n*=3,312), *n*=182 did not provide any data on the task; *n*=70 were omitted due to negative *d’* scores and/or not responding to over 50% of the trials, leaving a sample of *n*=3,242 (M=2.75, SD=0.81).

### Response inhibition

The Stop Signal Task (36) was used to assess response inhibition – the ability to prevent an ongoing motor response. The task consisted of 256 trials, which included a 4:1 ratio of trials without stop signals to trials with stop signals. Mean response times were calculated. An estimate of *stop signal reaction time* (SSRT) was calculated and used as the primary outcome as it is a reliable measure of inhibitory control, with shorter SSRT’s indicating faster inhibition. SSRT data were available for *n*=3,201 (M=258.60, SD=53.19).

### Emotion recognition

Emotion recognition was assessed using a six alternative forced choice (6AFC) emotion recognition task (37) comprising of 96 trials (16 for each emotion) which measures the ability to identify emotions in facial expressions. In each trial, participants were presented with a face displaying one of six emotions: anger, disgust, fear, happiness, sadness, or surprise. An overall measure of ER (the number of facial emotions accurately identified) was used as the primary outcome. ER data were available for *n*=3,368 (M=0.69, SD=0.08).

### Potential confounders

We identified confounders from established risk factors for cognitive functioning that could plausibly have a causal relationship with earlier binge drinking including income, maternal education, socioeconomic position, housing tenure, sex, and maternal smoking during first trimester in pregnancy. Two measures were included to control for cognitive function prior to alcohol initiation. Working memory at approximately 11 years and experience of a head injury/unconsciousness up to 11 years. Finally, measures of cigarette and cannabis use were collected at 4 timepoints between ages ∼14 and ∼16.5 years (up to the first assessment of binge drinking). Further information on all measures are presented in Supplementary material.

## Statistical methods

### Observational analyses

The observational analyses were conducted in two stages. First, longitudinal latent class analysis was used to derive trajectories of binge drinking for individuals having at least one measure of binge drinking (*n*=6,353). Starting with a single latent class, additional classes were added until model fit was optimised. See supplementary material for a description of model fit. Analyses were carried out using Mplus 8.1 (38).

Class membership was then related to covariates using the three-step method using the Bolck-Croon-Hagenaars (BCH) (39) method. The first stage estimated the latent class measurement model and saves the BCH weights. While, the second stage involved using these weights which reflect the measurement error of the latent class variable. Linear regression was used to examine the association between the continuous distal outcomes and latent class membership controlling for the confounding variables. Results are reported as unstandardized beta coefficients and 95% confidence intervals.

### Missing data

Missing data was dealt with in three steps. Of those invited to the age 24-year clinic (*n*=9,299), 6,353 (68%) participants provided self-report information on binge drinking on at least one timepoint between 16 and 23 years. Of these, *n*=3,755 (59%) had available information on all covariates. Next, multiple imputation was based on 3,155 (46%) participants who had information on at least one of the cognitive outcomes. The imputation model (based on 100 datasets) contained performance on all of the cognitive tasks, all measures of binge drinking, and potential confounding variables, as well as a number of auxiliary variables known to be related to missingness (e.g., substance use in early adolescence, parental financial difficulties, and other SES variables). Finally, inverse probability weighting was used where estimates of prevalence and associations were weighted to account for probabilities of non-response to attending the clinic. See Table S1 for a detailed description of attrition.

### Genetic analyses

Two-sample MR was used to test the hypothesised causal effect of alcohol use on cognitive functioning. The two-sample MR approach requires summary level data from two GWAS, enabling SNP-exposure and SNP-outcome effects to be derived from different data sources. As the genetic instrument for alcohol consumption we used the 99 conditionally independent genome-wide significant SNPs associated with ‘number of drinks per week’, identified by the GWAS & Sequencing Consortium of Alcohol and Nicotine use (GSCAN https://gscan.sph.umich.edu/) based on a sample of *n*=941,280. The 99 SNPs explain 2.5% of the variance in number of drinks per week (27). 87 of these SNPs were available in ALSPAC. As outcomes, we used GWAS conducted in ALSPAC for each of our three primary cognitive measures: i) working memory assessed using *d’ (n*=2,471); ii) response inhibition assessed using SSRT (*n*=2,446); and iii) emotion recognition assessed using total number of correctly recognised emotions (*n*=2,560). Further information is provided in the Supplementary material (Figures S4-S9). The main strength of using summary data from large GWAS consortia in two-sample MR is the increased statistical power. Analyses were performed using the TwoSampleMR R package, part of MR-Base (40). Power calculations conducted for one-sample MR analyses using mRnd (41) indicated that we had 80% power to detect an effect size of 0.335 for number of drinks per week using a sample size of n∼2,500 (participants with available genetic data).

It should be noted that neither the study nor the analysis plan were pre-registered on a publicly available platform, so the results should be considered exploratory.

## RESULTS

### Observational analyses

#### Patterns of binge drinking

The prevalence of both monthly and weekly binge drinking increased across time apart from a slight decrease at age 23 years (Table S2). There was good agreement that a four-class solution was adequate in explaining the heterogeneity in binge drinking based on increasing Bayesian Information Criterion (BIC) (42) and sample size adjusted Bayesian Information Criterion (SSABIC) (43) values in the five-class model and an LRT value of *p*=0.05. See Table S3 in the Supplementary Material for a comparison of model fit indices. The four-class solution (Figure 1) comprised patterns of binge drinking that were labelled as ‘low-risk’ (47.1%), ‘early-onset monthly’ (19.0%), ‘adult frequent’ (18.7%), and ‘early-onset frequent’ (19.0%). See Table S4 in the Supplementary Material for class validation. A detailed description of confounding factors associated with binge drinking class membership is provided in Table S5.

**Figure 1.**
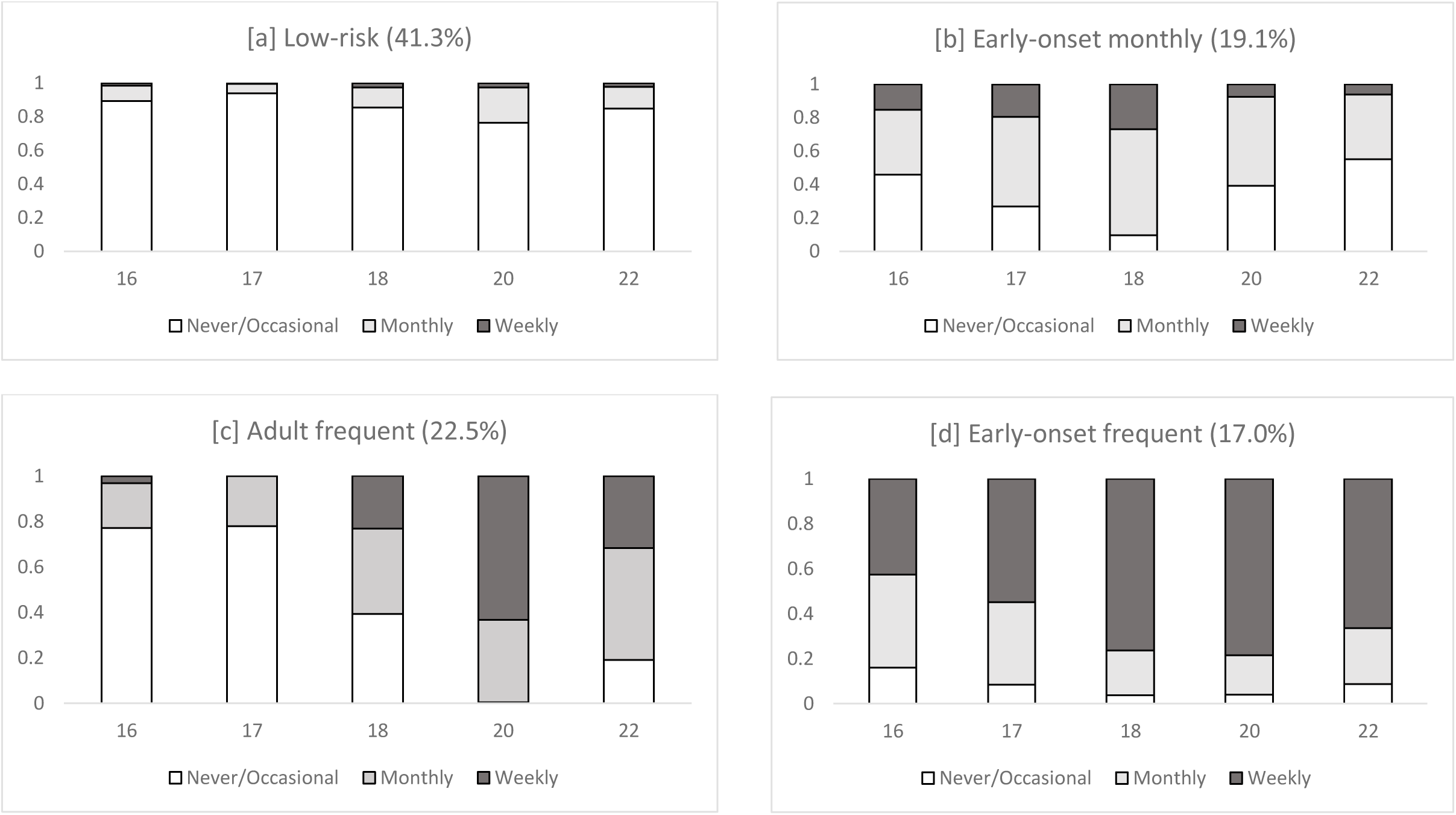
Distribution of binge drinking response across latent classes at each timepoint (*n*=6,353). Class proportions based on estimated posterior probability^1^. ^1^ Overall, the low-risk group reported a low probability of binge drinking across all measurement occasions; ‘early-onset monthly’ binge drinkers was mostly characterised by binge drinking in the earlier measurement occasions (but not later ones), ‘adult frequent’ binge drinkers were mostly characterised by binge drinking in the later measurement occasions (but not earlier ones), while ‘early-onset frequent’ binge drinking was mostly characterised by binge drinking across all timepoints.

#### Working memory – 2-back task

Table 1 presents unadjusted and adjusted associations between patterns of binge drinking from 16 to 23 years and working memory at age 24. There was little evidence to suggest an association between patterns of binge drinking and working memory performance in the fully adjusted models (‘early-onset monthly’: *b*=0.54, 95%CI=-1.92 to 0.82; ‘adult frequent’: *b*=0.03, 95%CI=-0.80 to 0.86; ‘early-onset frequent’: *b*=-0.42, 95%CI=-1.24 to 0.41). Furthermore, there was little evidence to suggest that patterns of binge drinking were associated with the secondary outcomes (i.e., number of hits and false alarms) (Table S6).

**Table 1.**
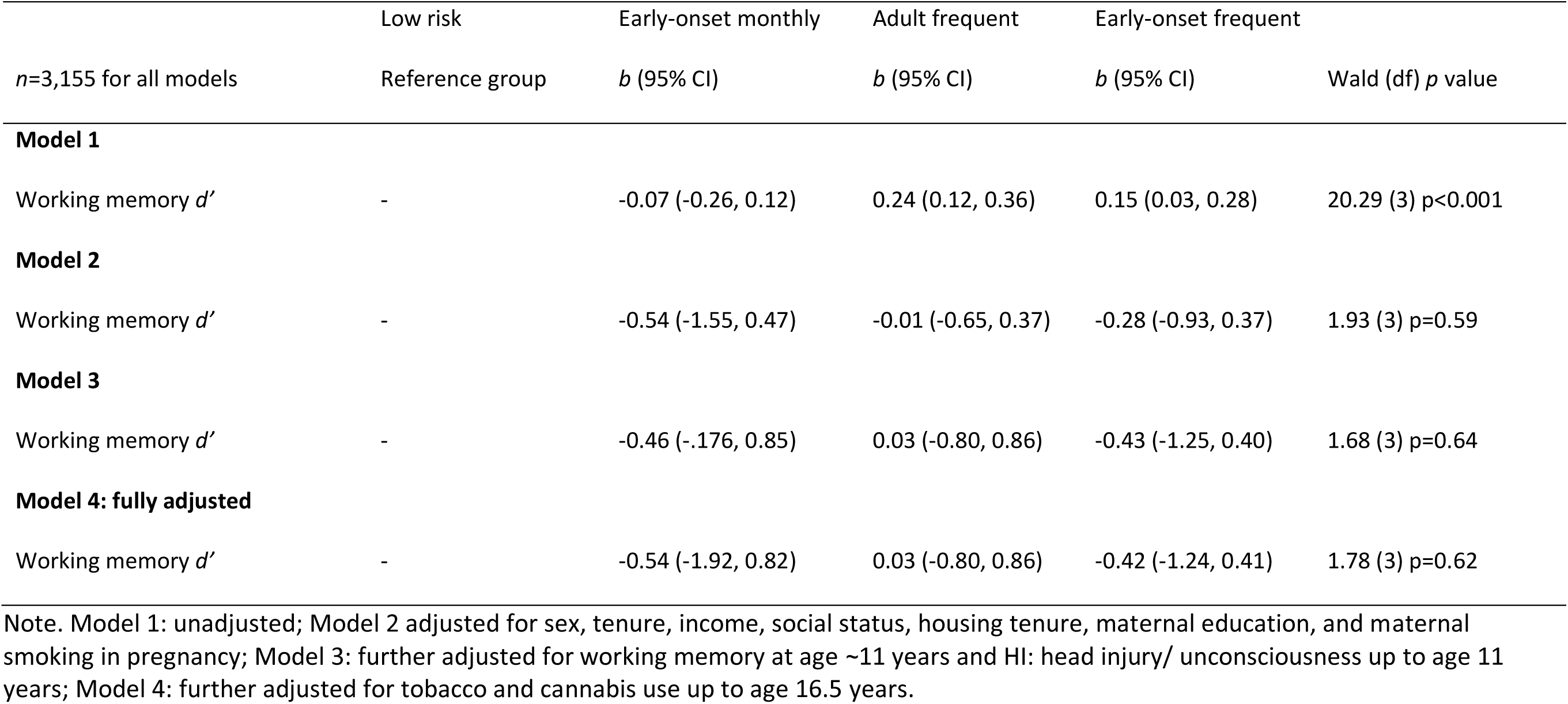
Patterns of binge drinking from 16 to 23 years and working memory at age 24 (higher *d’* scores reflect better performance)

[Table 1]

#### Response inhibition -stop signal task

Table 2 presents unadjusted and adjusted associations between patterns of binge drinking and an overall measure of response inhibition. There was little evidence to suggest an association between patterns of binge drinking and ability to inhibit responses in the fully adjusted models (‘early-onset monthly’: *b*=-3.9, 95%CI=-109.3 to 101.5; ‘adult frequent’: *b*=15.9, 95%CI=-38.2 to 69.9; ‘early-onset frequent’: *b*=31.9, 95%CI=-25.3 to 89.2). Furthermore, there was little evidence to suggest that patterns of binge drinking were associated with any of the secondary outcomes (i.e., Go reaction time, Go accuracy, and Stop accuracy) (Table S7).

**Table 2.** Patterns of binge drinking from 16 to 23 years and response inhibition at age 24 (shorter scores reflect faster reaction times)

|  | Low risk | Early-onset monthly | Adult frequent | Early-onset frequent |  |
| --- | --- | --- | --- | --- | --- |
| *n*=3,155 for all models | Reference group | *b* (95% CI) | *b* (95% CI) | *b* (95% CI) | Wald (df) *p* value |
| **Model 1** |  |  |  |  |  |
| Stop signal reaction time | - | 11.5 (-0.9, 23.8) | -8.7 (-16.8, -0.6) | -5.9 (14.0, 2.1) | 10.74 (3) *p*=0.01 |
| **Model 2** |  |  |  |  |  |
| Stop signal reaction time | - | 35.7 (-33.4, 104.8) | -12.3 (-54.9, 30.3) | 12.0 (-31.3, 55.2) | 2.13 (3) *p*=0.55 |
| **Model 3** |  |  |  |  |  |
| Stop signal reaction time | - | 5.6 (95.3, 106.4) | 15.7 (-38.3, 69.7) | 32.6 (-24.6, 89.8) | 1.39 (3) *p*=0.71 |
| **Model 4: fully adjusted** |  |  |  |  |  |
| Stop signal reaction time | - | -3.9 (-109.3, 101.5) | 15.9 (-38.2, 69.9) | 31.9 (-25.3, 89.2) | 1.35 (3) *p*=0.72 |

Note. Model 1: unadjusted; Model 2 adjusted for sex, tenure, income, social status, housing tenure, maternal education, and maternal smoking in pregnancy; Model 3: further adjusted for working memory at age ~11 years and HI: head injury/ unconsciousness up to age 11 years; Model 4: further adjusted for tobacco and cannabis use up to age 16.5years.

[Table 2]

#### Emotion recognition – 6AFC task

Table 3 presents unadjusted and adjusted associations between patterns of binge drinking and number of correctly identified emotions. There was little evidence to suggest an association between patterns of binge drinking and emotion recognition in the fully adjusted models (‘early-onset monthly’: *b*=0.01, 95%CI=-0.12 to 0.14; ‘adult frequent’: *b*=0.04, 95%CI=-0.04 to 0.13; ‘early-onset frequent’: *b*=0.02, 95%CI=-0.07 to 0.10). Furthermore, there was little evidence to suggest that patterns of binge drinking were associated with any of the secondary outcomes (i.e., anger, disgust, fear, happy, sad, and surprise) (Table S8).

**Table 3.**
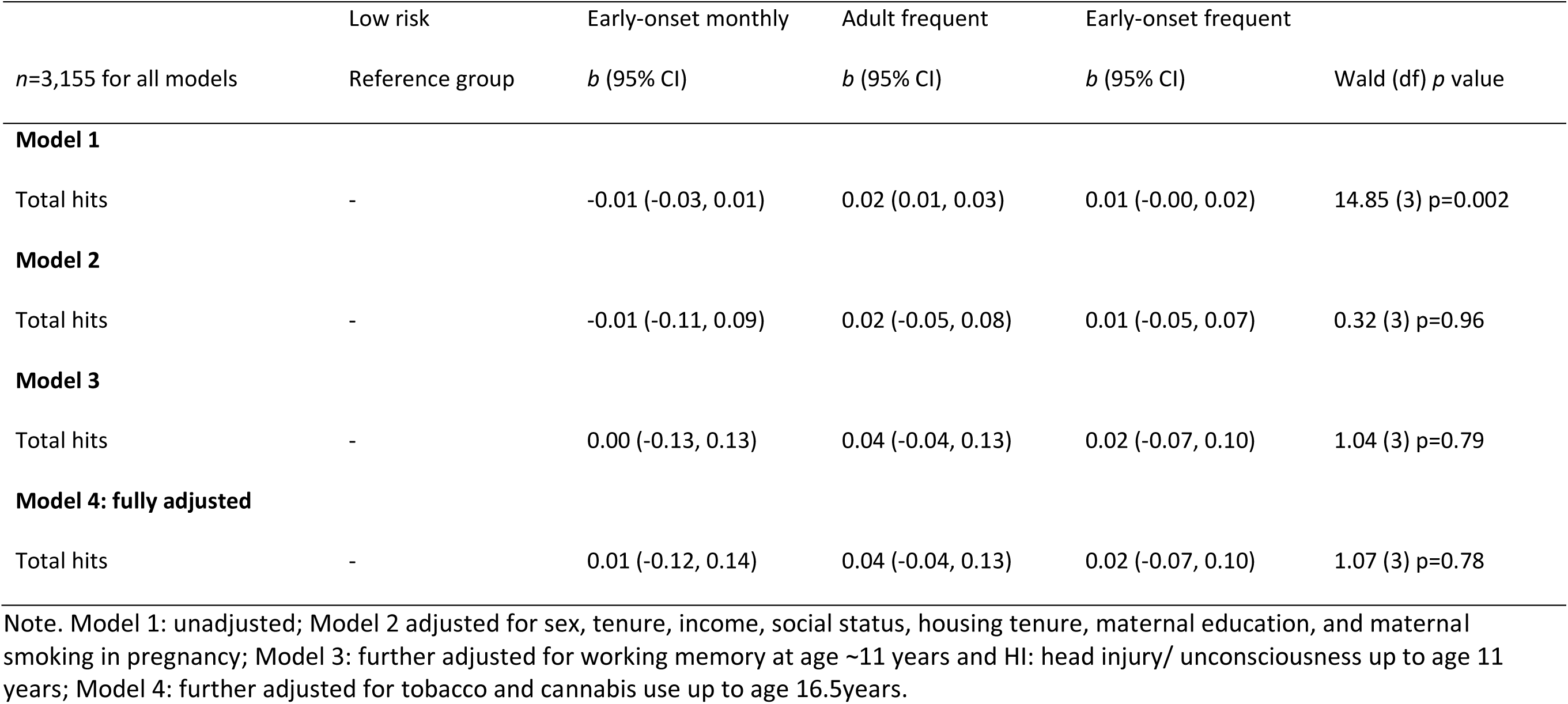
Patterns of binge drinking from 16 to 23 and emotion recognition at age 24 (higher scores reflect better performance)

#### Sensitivity analyses

Models using complete cases were included to assess the impact of missing data (Table S9). A latent growth model of the five repeated measures of binge drinking was conducted to examine the association with working memory, response inhibition, and emotion recognition while controlling for potential confounding variables (*n*=3,155) (Figure S3 and Table S10).

#### Genetic analyses

##### Mendelian randomisation

The two-sample MR method provides little evidence to suggest that alcohol use (SNPs associated with number of drinks per week) is a causal risk factor for deficits in cognitive functioning (Table 4). Focusing on the IVW estimate as the primary measure, SNPs associated with the number of alcoholic drinks per week were not associated with *d’* on the working memory task (*b*=0.285 95% CI=-0.42 to 0.99; *p*=0.43); SSRT on the response inhibition task (*b*=-0.321 95%CI=-1.04 to 0.39; *p*=0.38); or total hits in the emotion recognition task (*b*=0.028 95% CI=-0.55 to 0.61; *p*=0.93). Sensitivity analyses did not alter the main findings.

**Table 4.** Two-sample Mendelian randomization analyses of the effects of alcohol use on cognitive functioning.

| Exposure | Outcome | Method | N SNPs | Beta (95% CI) | P-value |
| --- | --- | --- | --- | --- | --- |
| **Drinks per week** | **Working memory** | Inverse-Variance Weighted | 87 | 0.285 (-0.42, 0.99) | 0.43 |
|  |  | MR Egger (SIMEX) | 87 | -0.473 (-1.70, 0.74) | 0.45 |
|  |  | Weighted Median | 87 | 0.408 (-0.50, 1.32) | 0.38 |
|  |  | Weighted Mode | 87 | 0.315 (-1.45, 2.08) | 0.32 |
| **Drinks per week** | **Response inhibition** | Inverse-Variance Weighted | 87 | -0.321 (-1.04, 0.39) | 0.38 |
|  |  | MR Egger (SIMEX) | 87 | -1.213 (-2.23, 2.20) | 0.29 |
|  |  | Weighted Median | 87 | -0.556 (-1.55, 0.43) | 0.27 |
|  |  | Weighted Mode | 87 | -0.689 (-2.49, 1.13) | 0.46 |
| **Drinks per week** | **Emotion recognition** | Inverse-Variance Weighted | 87 | 0.028 (-0.55, 0.61) | 0.93 |
|  |  | MR Egger (SIMEX) | 87 | 0.445 (-1.14, 2.03) | 0.58 |
|  |  | Weighted Median | 87 | -0.157 (-1.01, 0.69) | 0.72 |
|  |  | Weighted Mode | 87 | -0.180 (-1.78, 1.42) | 0.82 |

Note: SIMEX = simulation extrapolation. SIMEX-corrected estimates were used based on regression dilution $I_{2GX}$ for number of drinks per week values between 0.3 and 0.9. SIMEX-corrected estimates are unweighted.

## DISCUSSION

We found insufficient evidence to suggest an association between binge drinking between the ages of 16 and 23 and working memory, inhibition, and emotion recognition at age 24 using a combination of observational and genetic approaches. In the observational analyses, there was no evidence to suggest that binge drinking patterns identified in adolescence and early adulthood were associated with later measures of working memory, inhibition, and emotion recognition. While, the genetic analyses provided no clear causal evidence.

### Comparison with previous studies

Unlike the studies that have demonstrated a prospective association between binge drinking and cognitive functioning in adolescence e.g. (17–20), we found little evidence in support of this association. The contrast in findings could be due to a number of possibilities. First, our study used a large population sample incorporating data from over 3,000 individuals spanning maternal pregnancy to 24 years of age. Previous studies (19,20) used functional magnetic resonance imaging based on youths at high-risk for substance use disorders (*n*=40 for both studies), while the study by Peeters and colleagues (18) used a sample at high-risk for externalising problems (*n*=374 at baseline). Second, this study assessed cognitive functioning in early adulthood, a time when cognitions are thought to reach maturity (2–5), in comparison to all four studies who assessed cognitive functioning up to 19 years of age. Examining peak levels of cognitive functioning helps to reduce the possibility that cognitive functioning is influencing earlier alcohol use (i.e., reverse causation). Third, as alcohol use behaviours typically change over time (44), repeated measures of binge drinking were used in this study to capture heterogenous patterns across this sensitive period in comparison to our previous study which assessed alcohol use on one occasion (17). Finally, most of the previous studies (18–20) assessed cognitive functioning using different measures to the more widely used measures in this study (i.e., *N*-back task, Stop signal task, and 6AFC task).

Our findings support and extend those of Boelema and colleagues (45) who found insufficient evidence to suggest that alcohol use prospectively affected maturation of cognitive functions in a large prospective study of Dutch adolescents (46). First, Boelema and colleagues examined cognitive functioning across adolescence, while we were able to examine peak levels of cognitive functioning. Second, assessing binge drinking in young adulthood allowed us to capture heterogenous patterns during the sensitive period (i.e., going to University or in full-time paid employment). Finally, WM performance was measured in reaction times only, as opposed to the more comprehensive approach used in our study (i.e., number of hits, false alarms, and *d’*).

There are a number of differences with the findings from Kumari and colleagues (26). First, the observational study examines weekly alcohol consumption (and was dichotomised into ‘any versus none’ per week), compared to the repeated measures approach used in this study. Second, different cognitive outcome measures were used. Third, alcohol and cognition were assessed in mid-late adulthood (mean age across six studies ranged from 55 to 66 years) compared to adolescence/ young adulthood in this study. The MR analyses was based on a single SNP (rs1229984) as opposed to 87 SNPs in this study. The main disadvantage to using single SNPs is that statistical power may be low and an inability to separate horizontal from vertical pleiotropy (47).

### Limitations

First, the ALSPAC cohort suffers from attrition which reduces study power and is higher among the socially disadvantaged (48) (Table S1). We attempted to minimise the impact of attrition using sensitivity analyses. Missingness was related to having information on binge drinking, potential confounders and cognitive information. However, the pattern of results remained the same in the complete case (Table S9) and imputed analyses, suggesting that the pattern of missing data did not lead to biased effect estimates. Second, it is possible that both the observational and two-sample MR analyses are underpowered. For example, poor entropy (a measure of class separation) could indicate poor misclassification in the latent classes. Although binge drinking assessments spanned 7 years, it is possible that the use of a single item, with three possible response options, may diminish the ability to assess heterogenous drinking patterns. Misclassification of this kind (non-differential) when the exposure variable has more than two categories can bias the association in either direction (49) suggesting that true underlying associations could be stronger or weaker than we observed. Although this is likely, we are unable to validate these classes with an alcohol biomarker as an adequate biomarker for binge drinking in adolescents/young adulthood is not available in ALSPAC. Patterns of binge drinking were however shown to have a dose-response association with a later AUDIT-consumption measure, assessed at 24 years of age (Table S4).

Third, including yearly binge drinking assessments would have helped class formation, however given the pattern of results, it is plausible that they would not change the pattern of results. Fourth, although it is possible to either under-or over-estimate drinking behaviour using self-reported data, participants completed questionnaires individually and were assured of their anonymity. Fifth, different measures of alcohol use for the observational and

MR analyses were used. Along with deriving latent classes of binge drinking, we used the largest GWAS consortia (GSCAN) which has identified 87 genetic instruments for ‘number of alcohol drinks per week’ which is a continuous measure. To our knowledge it is not currently possible to use a nominal exposure (as was used in the observational analyses) and consequently the effect sizes are not directly comparable.

Sixth, as we examined one potential causal pathway, it is possible that the association could work in the opposite direction, that is, impairments in cognitive functioning may precede (and increase the risk of developing) alcohol problems (18,23). We were however able to include a number of measures to maximise the robustness of our findings: (i) ascertaining the time order of exposures and outcomes; (ii) controlling for premorbid working memory function and brain insults prior to the onset of alcohol use; and (iii) a number of relevant confounders were included to help reduce the possibility of residual confounding. It is possible that a common risk factor is influencing both binge drinking and deficits in cognition, however the two-sample MR analyses helps to protect against this possibility by minimising bias from reverse causation and residual confounding. Seventh, genetic variants were based on number of drinks per week, whereas the observational analyses used frequency of binge drinking. Although not directly comparable, there was evidence of a dose response relationship between binge drinking patterns and the AUDIT-C measure, which taps into quantity and frequency (see Supplementary material).

Finally, the main limitation of two-sample MR is that the quality of the pooled results in the GWAS consortia is dependent on the individual studies. Another limitation is that the same sample may contribute to both GWAS (i.e., GWAS for exposure and outcome) which was the case in the current study as ALSPAC was in both the exposure and outcome. This will bias the MR estimate towards the observed estimate. However, as the MR found no clear evidence for an effect, this suggests it was not biased by overlapping samples. See Lawlor et al. (25) for a more comprehensive description of limitations associated with MR studies.

### Implications and Conclusions

In order to rule out the possibility of deficient cognitive functioning preceding binge drinking in adolescence, future research should use an equally robust approach to examine the alternate hypothesis. We found insufficient evidence to suggest an association between binge drinking between the ages of 16 and 23 and cognitive deficits at age 24 using a combination of observational and genetic approaches, although both approaches are likely to be underpowered. Future studies should use larger observational samples and meta-analyses of related cognitive measures in GWAS to help to increase power.

## Data Availability

We used data from the Avon Longitudinal Study of Parents and Children (ALSPAC), an ongoing population-based study that contains a wide range of phenotypic and environmental measures, genetic information and linkage to health and administrative records.

http://www.bristol.ac.uk/alspac/researchers/our-data/

http://www.bris.ac.uk/alspac

## Acknowledgements

The work was undertaken with the support of the MRC and Alcohol Research UK (grant number MR/L022206/1). We acknowledge also support from The Centre for the Development and Evaluation of Complex Interventions for Public Health Improvement (DECIPHer); a UKCRC Public Health Research Centre of Excellence (joint funding (grant number MR/KO232331/1) from the British Heart Foundation, Cancer Research UK, Economic and Social Research Council, Medical Research Council, the Welsh Government and the Wellcome Trust, under the auspices of the UK Clinical Research Collaboration); and the NIHR School of Public Health Research. Support was also provided by the UK Medical Research Council Integrative Epidemiology Unit at the University of Bristol (MM_UU_00011/7). LM, RW, SS, and MRM are members of the UK Centre for Tobacco and Alcohol Studies, a UKCRC Public Health Research Centre of Excellence. LM, REW, SS, and MRM are supported by the NIHR Biomedical Research Centre at the University Hospitals Bristol NHS Foundation Trust and the University of Bristol. The views expressed in this publication are those of the authors and not necessarily those of the NHS, the National Institute for Health Research or the Department of Health and Social Care. GWAS data were generated by Sample Logistics and Genotyping Facilities at Wellcome Sanger Institute and LabCorp (Laboratory Corporation of America) using support from 23andMe. The UK Medical Research Council and Wellcome (Grant ref: 102215/2/13/2) and the University of Bristol provide core support for ALSPAC. This publication is the work of the authors and LM and MRM will serve as guarantors for the contents of this paper. A comprehensive list of grants funding is available on the website (http://www.bristol.ac.uk/alspac/external/documents/grant-acknowledgements.pdf). We are extremely grateful to all the families who took part in this study, the midwives for their help in recruiting them, and the whole ALSPAC team, which includes interviewers, computer and laboratory technicians, clerical workers, research scientists, volunteers, managers, receptionists and nurses. This publication is the work of the authors who will serve as guarantors for the contents of this paper.

